# A mixed-methods study comparing digitized versus paper-based tools during the provision of sexual and reproductive health services for young women in Ethiopia

**DOI:** 10.64898/2026.05.12.26353066

**Authors:** Bekele Belayihun, Meghan Cutherell, Abednego Musau, Fana Abay, Alexis Coppola

## Abstract

Adolescent girls and young women (AGYW) in Ethiopia face persistent barriers to accessing quality sexual and reproductive health (SRH) services, including limited information, stigma, and lack of youth-responsive care. This study aimed to compare the efficacy of digitized versus paper-based counselling tools within an intervention designed to address behavioral and structural barriers contributing to low contraceptive use among AGYW, by reframing contraception as a tool to achieve their life goals.

The study employed a cross-sectional mixed-methods design, including client exit interviews with 302 AGYW, key informant interviews with 18 Health Extension Workers (HEWs), secondary analysis of service delivery data from DHIS2, and costing data from program records. Quantitative data were analyzed using descriptive statistics and chi-square tests. Qualitative data were thematically analyzed. Digital counselling was significantly associated with higher MII Plus scores (93% vs. 73.8%, p=0.001), client knowledge of side effects, and confidence in discussing and managing contraception. Clients rated paper-based tools as easier to understand, but digital tools enhanced comprehension, goal-setting, and integration of financial planning and reproductive health concepts. HEWs reported improved consistency in counselling, better referrals, and operational efficiencies with digital tools. Challenges included device glitches, limited connectivity, and variable digital literacy, often requiring concurrent use of paper and digital tools.

This study shows that transitioning from paper-based to digital counselling tools improved service quality, client engagement, and informed contraceptive decision-making. Higher MII Plus scores and positive client experiences indicate more standardized, participatory, and respectful counselling. Providers reported operational benefits, including easier counselling and improved data management, though productivity gains were limited. Implementation challenges highlight the need for context-sensitive strategies, ongoing training, and supportive supervision during digital integration. Importantly, the findings suggest that digital tools can improve how services are delivered (quality and consistency), even when service volume remains stable.

**AUTHOR SUMMARY:** This study looked at how to improve access to family planning services for adolescent girls and young women in Ethiopia, who often face stigma, lack of information, and unfriendly healthcare services. We compared two ways of providing family planning counselling: a traditional paper-based guide and a digital tool (implemented on a tablet or phone). We gathered information from over 300 young women and healthcare workers and analyzed service records to compare differences between the two approaches. We found that the digital tool resulted in improved quality of counselling. Young women better understood their contraceptive options, including side effects, and felt more confident making decisions. The digital tool also helped to better connect family planning to personal goals, like education or financial planning. However, many participants found the paper tool easier to follow. Healthcare workers reported that the digital tool made their work more consistent and efficient, but they also faced problems like poor internet, device malfunctions, and varying comfort with technology. Sometimes they had to use both the digital and paper tool together. Overall, the digital tool showed clear benefits, but challenges mean they should be introduced carefully, with proper training and support to ensure they work effectively in real-world settings.

## INTRODUCTION

### Adolescent Sexual and Reproductive Health in Ethiopia

Adolescent and youth sexual and reproductive health (AYSRH) in Ethiopia has seen significant improvements over the past few decades, driven by expanded national programs and increased attention to the needs of adolescents and youth.^1^ Ethiopia has one of the largest youth populations in Africa – about 32 million people aged 10-24 – making AYSRH a critical priority.^2^ Ethiopian adolescents face a high burden of sexual and reproductive health (SRH) issues, ranging from early marriage and sexual debut to rapid and repeat pregnancies, unsafe abortion, sexually transmitted infections, and harmful traditional practices.^3^ Though demonstrable progress has been made in improving outcomes across these areas in recent decades – such as reductions in child marriage and adolescent fertility – gains remain uneven across regions and population groups.^1,4^

Early marriage continues to be a key driver of AYSRH risks in Ethiopia, through its close link to early sexual initiation and childbearing. In the 2024 demographic health survey, 40% of women aged 20-24 are were married before age 18 and 13% of adolescent girls (15-19) had begun childbearing, with large disparities between rural (15-16%) and urban (6-7%) adolescent girls.^4^ Use of modern contraception among married adolescent girls increased from 32% to 36% between 2019 and 2024, but one in five married girls still has an unmet need for contraception.^4^ Overall, adolescents – especially rural and disadvantaged girls – remain a high-risk population requiring targeted interventions.

### Digitization

Enabled by rapid growth in mobile phone penetration, digital health tools have transformed health promotion and service delivery globally and have created new opportunities to deliver health services and promote healthy behaviors. Digital technologies are increasingly being used to address persistent barriers to contraceptive uptake – including stigma, lack of information, limited youth-friendly services, and geographic inaccessibility.^5^ Mobile or tablet-based tools such as interactive, SMS, or IVR applications can be used to provide contraceptive information and counselling, appointment reminders, or guidance on contraceptive method selection.^5^

Digital tools can be complementary solutions that expand access to SRH information and services, especially for AGYW who face the greatest unmet need. MHealth interventions have demonstrated effectiveness in improving uptake of SRH services among adolescents, particularly contraceptive use.^6^ Those tools which are most successful are grounded in evidence and apply strategic behavior change techniques. The effectiveness of these tools also depends on their interactivity and integration with health systems.^7^

### Program Description

The Smart Start intervention was designed to address the behavioral and structural barriers that drive low contraceptive use among married adolescent girls in Ethiopia by reframing contraception as a tool for achieving life goals. Through structured conversations about financial planning, future aspirations, and the economic implications of early childbearing, Smart Start supports adolescent girls and young couples in making informed decisions about the timing and spacing of pregnancies.^8^ Through Smart Start, HEWs provide counselling using a tool that combines compelling visuals and analogies that support girls and couples to envision their goals for the future and understand how delaying and spacing births is relevant for the future health and prosperity of their family.^9^

Beginning in 2019, the Ethiopian Federal Ministry of Health (MOH), with technical support from Population Services International (PSI) Ethiopia, took leadership to scale Smart Start to more than 9,500 health posts across Oromia, Amhara, Tigray, Southwest, Central, South, Sidama, Somali, and Afar regions. Subsequently, in November 2023 the MOH launched a proposal for national scale-up of Smart Start, aiming to expand the intervention to all health facilities across Ethiopia.

As Smart Start scaled, several operational challenges emerged. Paper-based counselling materials were difficult to standardize across large geographic areas, supervision and quality monitoring were limited, and capturing counselling data for program learning was resource-intensive. These challenges became even more pronounced during the COVID-19 pandemic, which disrupted community engagement, reduced health facility visits, and limited opportunities for in-person counselling sessions.

To mitigate these challenges, PSI Ethiopia began exploring digital delivery mechanisms for Smart Start counselling. The initial digitization effort focused on transforming the paper-based counselling tool into a multimedia digital format. The digital version of the Smart Start counselling tool incorporated structured counselling flows, visual storytelling elements, and interactive prompts that guided HEWs through key discussion points related to family planning, gender-based violence prevention, and health service referrals. Beginning in May 2024, PSI Ethiopia collaborated closely with the MOH to integrate the digitized Smart Start tools directly into the national electronic Community Health Information System (eCHIS).

The objective of this research study was to compare, through triangulation of mixed methods data, the benefits and drawbacks of these two counselling modalities. The study aimed to provide comparison of these two approaches across a variety of thematic areas, including quality of service delivery, client experience, informed decision-making, healthcare worker experiences, system performance, site-level productivity, and cost-efficiency.

## RESULTS

### Client experience and quality of care

We surveyed 302 AGYW through client exit interviews (CEIs) (digital group = 114, paper-based group = 188). Clients were more likely to be in the 20–24-year-old age cohort, out of school, and residing in a rural area (Table 1). The majority (97%) were currently living with their spouse. Three-quarters of participants had already given birth. Statistically significant differences were observed between clients from digital sites and paper-based sites in location (with digital sites more rural) and religion (more participants in digital sites selecting ‘other’ for their religion).

**Table 1:** Participant socio-demographic characteristics from client exit interviews in sites using digital or paper-based Smart Start tool.

| Variable | Categories | Overall (N=302) | Digital group (n=114) | Paper-based group (n=188) | p-value |
| --- | --- | --- | --- | --- | --- |
| Location | Urban | 100 (33.1%) | 26 (22.8%) | 74 (39.4%) | 0.003 |
|  | Rural | 202 (66.9%) | 88 (77.2%) | 114 (60.6%) |  |
| Age (in years) | 15-19 | 87 (28.8%) | 32 (28.1%) | 55 (29.3%) | 0.826 |
|  | 20-24 | 215 (71.2%) | 82 (71.9%) | 133 (70.7%) |  |
| Living with spouse | Yes | 292 (96.7%) | 108 (94.7%) | 184 (97.9%) | 0.140 |
|  | No | 10 (3.3%) | 6 (5.3%) | 4 (2.1%) |  |
| Currently in school | Yes | 20 (6.6%) | 9 (7.9%) | 11 (5.9%) | 0.489 |
|  | No | 282 (93.4%) | 105 (92.1%) | 177 (94.2%) |  |
| Highest level of education attained | None | 55 (18.2%) | 28 (24.6%) | 27 (14.4%) | 0.061 |
|  | Primary | 142 (47.0%) | 46 (40.4%) | 96 (51.1%) |  |
|  | Secondary | 92 (30.5%) | 33 (29.0%) | 59 (31.4%) |  |
|  | Above secondary | 13 (4.3%) | 7 (6.1%) | 6 (3.2%) |  |
| Religion | Orthodox | 82 (27.2%) | 37 (32.5%) | 45 (23.9%) | <0.001 |
|  | Muslim | 171 (56.6%) | 28 (24.6%) | 143 (76.1%) |  |
|  | Others | 49 (16.2%) | 49 (43.0%) | 0 |  |
| Ever given birth | Yes | 229 (75.8%) | 86 (75.4%) | 143 (76.1%) | 0.128 |
|  | No | 73 (24.2%) | 28 (24.6%) | 45 (23.9%) |  |
| Heard of Smart Smart before visit | Yes | 242 (80.1%) | 92 (80.7%) | 150 (79.8%) | 0.847 |
|  | No | 60 (19.9%) | 22 (19.3%) | 38 (20.2%) |  |

Across both groups most clients adopted a contraceptive method during the visit, with only 10% receiving counselling without adopting a method (10% received counselling alone for the digital group compared to 8% for the paper-based group for which an additional 2% received a removal for an existing method) (Figure 1). Conversion rates (the proportion of clients who chose to receive a contraceptive method during the visit) was 89.8% overall without a statistically significant difference between the two groups (90.4% for the paper-based group and 88.8% for the digital group, *p*=0.705). Dominated by injectables (DMPA), implants, and pills, the method mix was significantly different between groups (*p*=0.023). Clients in the digital group had a higher frequency of using pills compared to those in the paper-based group who were more likely to adopt an injectable method.

**Figure 1:**
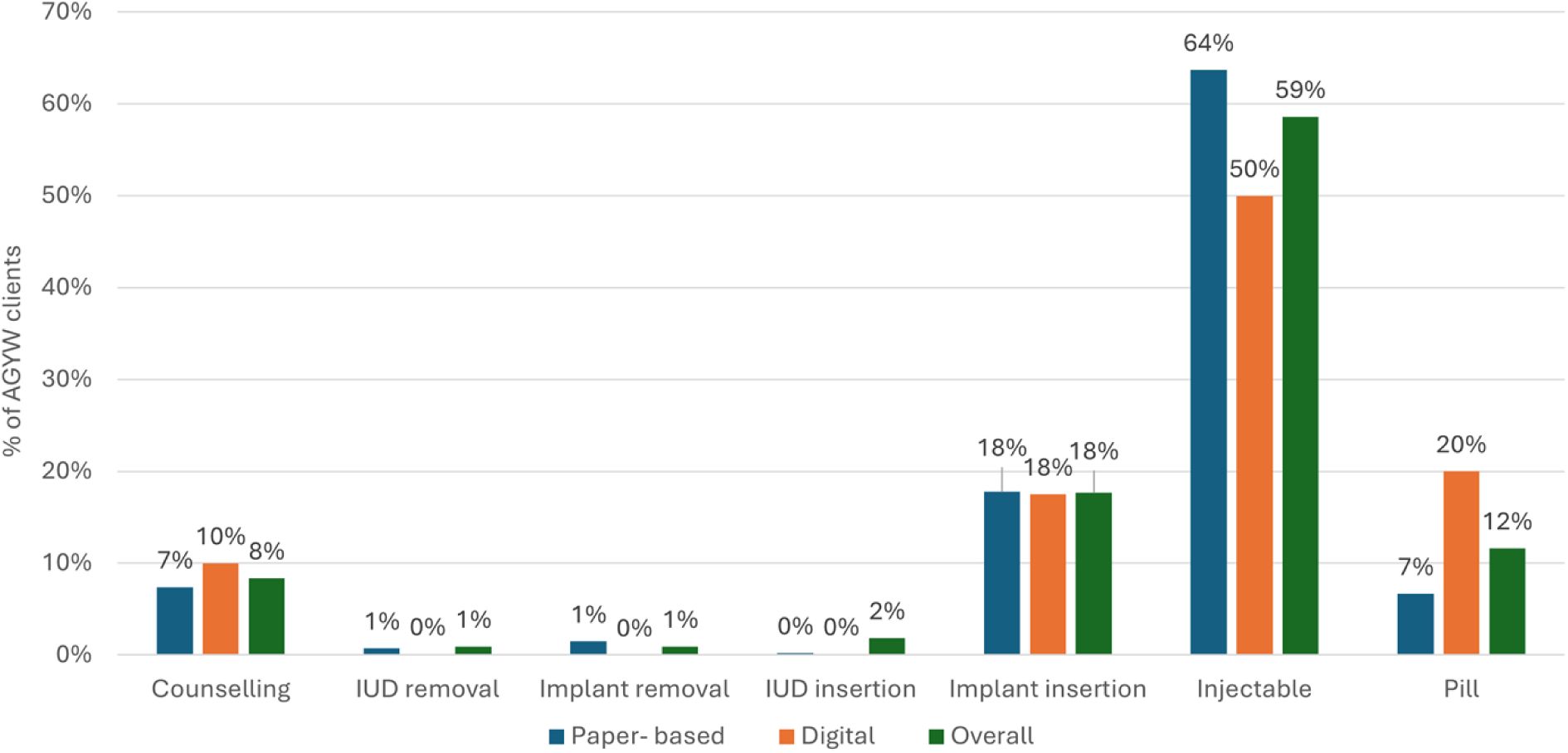
Counselling outcomes and method mix comparison from client exit interviews in sites using digital or paper-based Smart Start tool.

93% of participants in the digital group responded ‘yes’ to all four questions on the MII Plus compared to 73.8% of participants in the paper-based group (*χ*=10.663, *p*=0.001) (Table 2). Overall, 80.8% of participants in both groups responded ‘yes’ to all the four MII Plus questions.

**Table 2:** Responses to the Method Information Index (MII) Plus questions from client exit interviews in sites using digital or paper-based Smart Start tool.

| Individual questions to MII Plus* | Digital group (n= 71) | Paper-based group (n=122) | P value |
| --- | --- | --- | --- |
| During your counselling session, were you informed about other methods of contraception? | 70 (98.6%) | 119 (97.5%) | 0.621 |
| Were you informed about possible side effects or problems that you might experience when using the method? | 66 (93.0%) | 101 (82.8%) | 0.046 |
| Were you told what to do if you experience any side effects or problems? | 66 (93.0%) | 95 (77.9%) | 0.007 |
| Were you told about the possibility of switching to another method if the method you selected was not suitable? | 67 (94.4%) | 112 (91.8%) | 0.508 |
| **‘Yes’ to all four questions** | **66 (93.0%)** | **90 (73.8%)** | **0.001** |

\*Analysis includes 193 clients who adopted a method during the visit (excludes AGYW who did not adopt a method)

Clients in the digital group were more likely to report higher confidence in discussing contraceptive use as well as greater awareness of and improved preparedness to manage side effects (Table 3). This difference was statistically significant on several statements, including confidence in discussing method use with someone else (*p*=0.046), awareness of side effects of methods (p=0.018), and preparedness to handle these side effects (*p*=0.016). Perceptions of the paper-based tool were generally more positive than those of the digitalized tool. Significantly more clients in the paper-based group agreed that the tool was ‘good’ (*p*=0.011) and easy to understand (*p*=0.025) and that they liked the images in the tool (*p*=0.026).

**Table 3:** Client knowledge and confidence with contraceptives and their perception of the counselling tool from client exit interviews in sites using digital or paper-based Smart Start tool.

|  | Digital group<br>(n=114) | Paper-based<br>group<br>(n=188) | *p*-value |
| --- | --- | --- | --- |
| **Knowledge and confidence with contraceptives** |  |  |  |
| My knowledge about the methods to avoid or delay pregnancy has increased | 96 (86.5%) | 151 (83.0%) | 0.422 |
| I am more confident to have a discussion with someone about the methods to avoid or delay pregnancy | 108 (94.7%) | 165 (87.8%) | 0.046 |
| I am confident that I can make a choice about which methods to avoid or delay pregnancy I will use | 111 (94.4%) | 174 (92.6%) | 0.079 |
| I am more confident to have a discussion with my husband about the methods to avoid or delay pregnancy | 108 (94.7%) | 169 (89.9%) | 0.139 |
| I am less anxious about the method I will use to avoid or delay pregnancy | 70 (61.4%) | 112 (59.6%) | 0.753 |
| I am aware of the side effects or problems I might encounter when using a method to avoid or delay pregnancy | 97 (85.1%) | 138 (73.4%) | 0.018 |
| I am prepared to handle the side effects or problems I might encounter when using a method to avoid or delay pregnancy | 102 (89.5%) | 148 (78.7%) | 0.016 |
| **Perception of the counselling tool** |  |  |  |
| The tool which the provider used was good | 88 (77.2%) | 166 (88.3%) | 0.011 |
| The tool which the provider used was interesting | 88 (77.2%) | 156 (83.0%) | 0.216 |
| The tool which the provider used during the counselling was easy to understand | 80 (70.2%) | 153 (81.4%) | 0.025 |
| I like the images in the tool which the provider used during the counselling | 83 (79.5%) | 157 (83.5%) | 0.026 |
| The provider took a long time going through the tool which he/she used during the counselling | 50 (43.9%) | 94 (50.0%) | 0.300 |
| I gained a lot of information from the tool which the provider used during the counselling | 89 (78.1%) | 159 (84.6%) | 0.153 |
| The images in the tool which the provider used during the counselling clearly show how my husband and I can develop our lives | 84 (73.7%) | 152 (80.9%) | 0.144 |

93% of clients in the digital group rated their counselling experience as ‘good’ compared with 85.1% in the paper-based group (Table 4). The difference was weakly statistically significant (*p*=0.041). All clients across both groups reported that doctors, nurses, and other staff at the facility treated them with respect. There were some differences between the two groups in client perceptions of differential treatment according to personal attributes. More clients in the paper-based group reported that they were treated differently because of any personal attribute (*p*<0.001), their marital status (*p*=0.001), and how many children they had (*p*=0.011). The paper-based group was also more likely to indicate an absence of auditory and visual privacy.

**Table 4:** Client quality of care perceptions from client exit interviews in sites using digital or paper-based Smart Start tool.

|  | Digital group (n=114) | Paper-based group (n=188) | *p*- value |
| --- | --- | --- | --- |
| Comfort, respect, and perceived treatment |  |  |  |
| Did the doctors, nurses, or other staff at the facility treat you with respect as a person? | 114 (100%) | 188 (100%) | - |
| Did the HEW look at you or touch you in a way that made you feel uncomfortable? | 5 (4.4%) | 18 (9.6%) | 0.099 |
| During your time in the health facility, would you say you were treated differently because of any personal attribute like your age? | 9 (7.9%) | 46 (24.5%) | <0.001 |
| Were you treated differently because of your marital status? | 6 (5.3%) | 37 (19.7%) | 0.001 |
| Were you treated differently because of how many children you have? | 3 (2.6%) | 10 (10.6%) | 0.011 |
| Did you feel comfortable to ask questions during this visit? | 101 (88.6%) | 166 (76.6%) | 0.010 |
| Privacy and confidentiality |  |  |  |
| When meeting with the HEW during your visits, do you think other clients could hear what you and the HEW discussed? | 8 (7.05%) | 38 (20.2%) | 0.002 |
| Did you feel like your health information will be kept confidential at this facility? | 96 (84.2%) | 140 (78.7%) | 0.241 |
| When meeting with the HEW during your visits, do you think other clients could see you? | 13 (11.4%) | 49 (26.1%) | 0.002 |
| Counselling quality and information provision |  |  |  |
| Did you feel the doctors, nurses, and other staff at the facility clearly explained things to you? | 104 (91.2%) | 170 (90.4%) | 0.816 |
| Did the HEW let you say what mattered to you about your method?* | 75 (94.8%) | 126 (93.3%) | 0.905 |
| Did the HEW take your preferences about family planning methods seriously?* | 77 (96.3%) | 125 (92.6%) | 0.277 |
| Did the HEW you saw today give you enough information to make the best decision about your method?* | 71 (88.8%) | 127 (94.1%) | 0.162 |
| Did the HEW pressure you to use the method they wanted you to use?* | 8 (10.0%) | 34 (25.1%) | 0.007 |
| Did you receive information about what to do if you wanted to stop using a method?* | 77 (96.3%) | 114 (84.4%) | 0.008 |
| During your consultation today, did the HEW tell you about reasons why you would need to return? | 108 (94.7%) | 172 (91.5%) | 0.292 |
| Did you feel you had enough time during the consultation with the HEW to discuss and address your needs? | 108 (94.7%) | 172 (91.5%) | 0.292 |

*Table caption: \*Questions were asked to 215 AGYW (digital group = 80; paper-based group = 135) primarily visiting the facility to receive an FP method*

A greater proportion of clients in the paper-based group indicated that their life goals changed after their visit (58.3% in the digital group vs. 71.0% in the paper-based group, *p*=0.037), however clients in the digital group cited a greater variety of goals compared to those in the paper-based group (Figure 2). Clients in the digital group were more likely to indicate that they had financial (75% vs. 51%, p<0.001), educational (29% vs. 10%, p<0.001), and social relationship (25% vs. 12%, p=0.008) goals compared to peers in the paper-based group. Though clients in the paper-based group were more likely to say they had health-related and career goals, these differences were not statistically significant.

**Figure 2:**
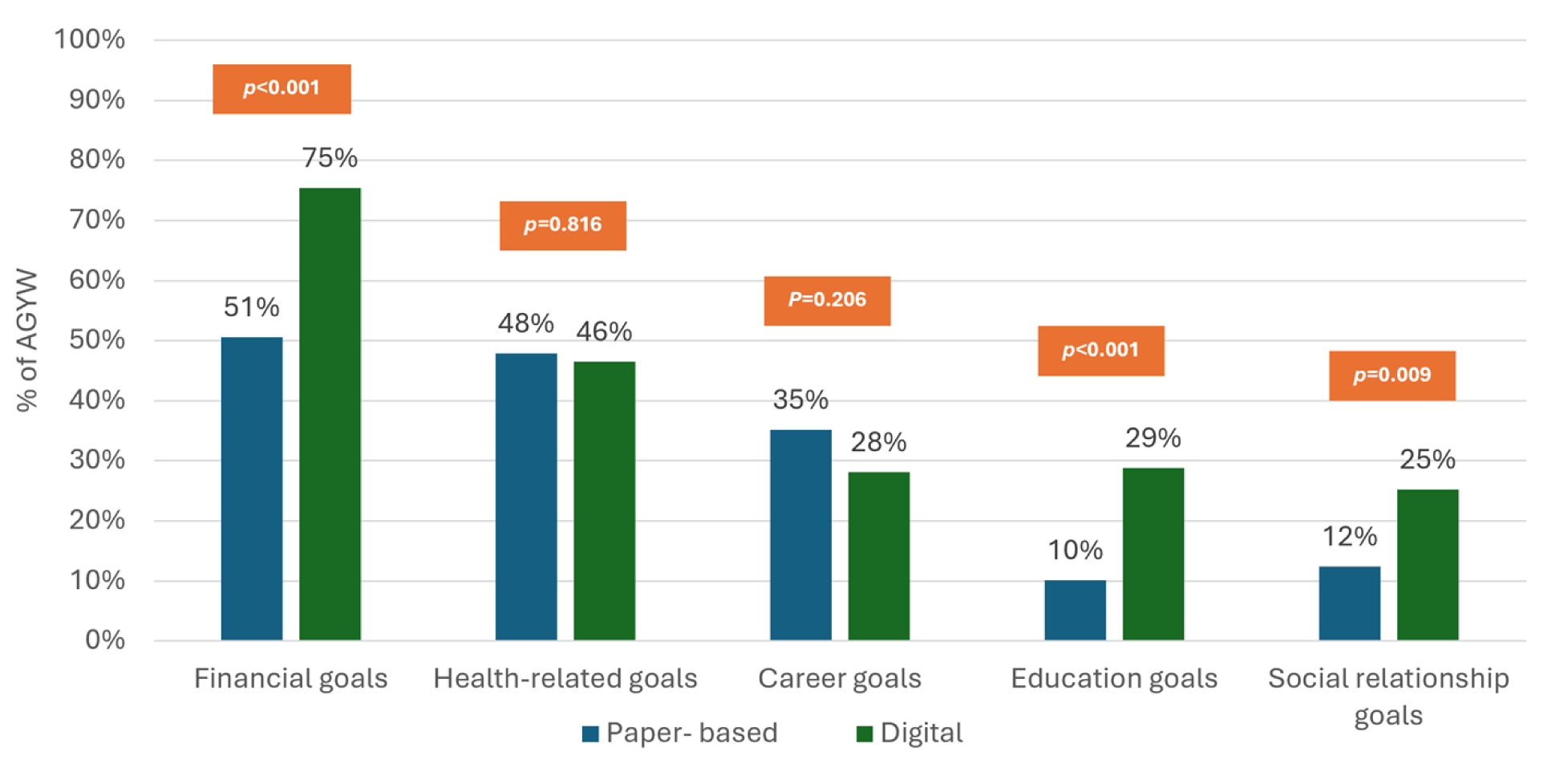
Comparison of AGYW stating they have goals across various thematic areas at the end of the visit from client exit interviews in sites using digital or paper-based Smart Start tool. Figure footnote: each goal type represents the proportion of clients who said that they had goals of this nature.

### Provider and operational experience

Within the qualitative sample, most participants had been using the paper-based tool for a period of 2-3 years prior to switching to the digital tool. Participants validated the superiority of the Smart Start paper-based tool compared to other counselling tools used within the health system. The tool was cited as being more visually appealing and innovative with its linkages between financial planning and contraception. The tool gave HEWs appropriate, targeted messages to reach clientele with high need and enhanced client comprehension of SRH concepts through its visualizations.

> *“It is very effective because based on the counsel which they took they prepared their life plan (business, saving, delaying pregnancy, planning childbearing time). For instance, one of my clients based on her plan now is finishing building a house. Others also started their own business and savings. The picture which represents the amount of food taken by one child per year is very interesting for the clients and it changes the attitude of the clients. Hence, they show willingness to delay pregnancy after they understand what the pictures are telling. Now a lot of clients are using different types of methods to delay pregnancy.”* – HEW, Chencha Zuria Woreda

At the time that the interviews were conducted, most HEWs had been using the digital tool for around six months and cited several of its advantages. HEWs indicated that the digital tool expanded on information that was available in the paper-based guide – particularly interaction around goal setting and financial planning concepts – and felt that the digital content was more accessible for client understanding (Table 5). HEWs also felt that the digital tool facilitated greater consistency in client-led contraceptive counselling approaches, supporting AGYW to have greater understanding of their method options, leading to higher uptake of long-acting methods and greater client preparedness for what they might experience with their method use. HEWs reported that the tool promoted greater integration of their different health area portfolios and enabled more effective referrals for clients with health needs outside of SRH. Additionally, they cited improved reporting with the digital tool – particularly capturing of client demographic information or follow-up appointments. In some cases, HEWs indicated that these operational efficiencies decreased effort needed to counsel clients, thereby reducing their overall workload. However, these improvements did not consistently translate into increased service volume, highlighting that digital tools improve quality and workflow even in the absence of these additional outputs.

**Table 5:**
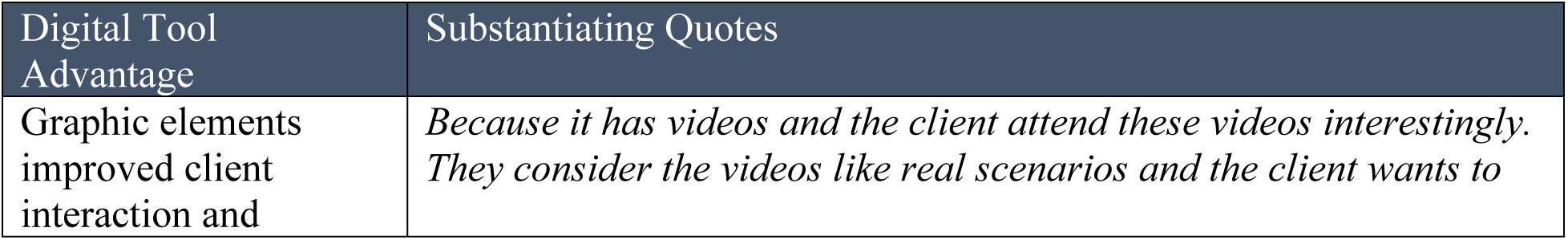
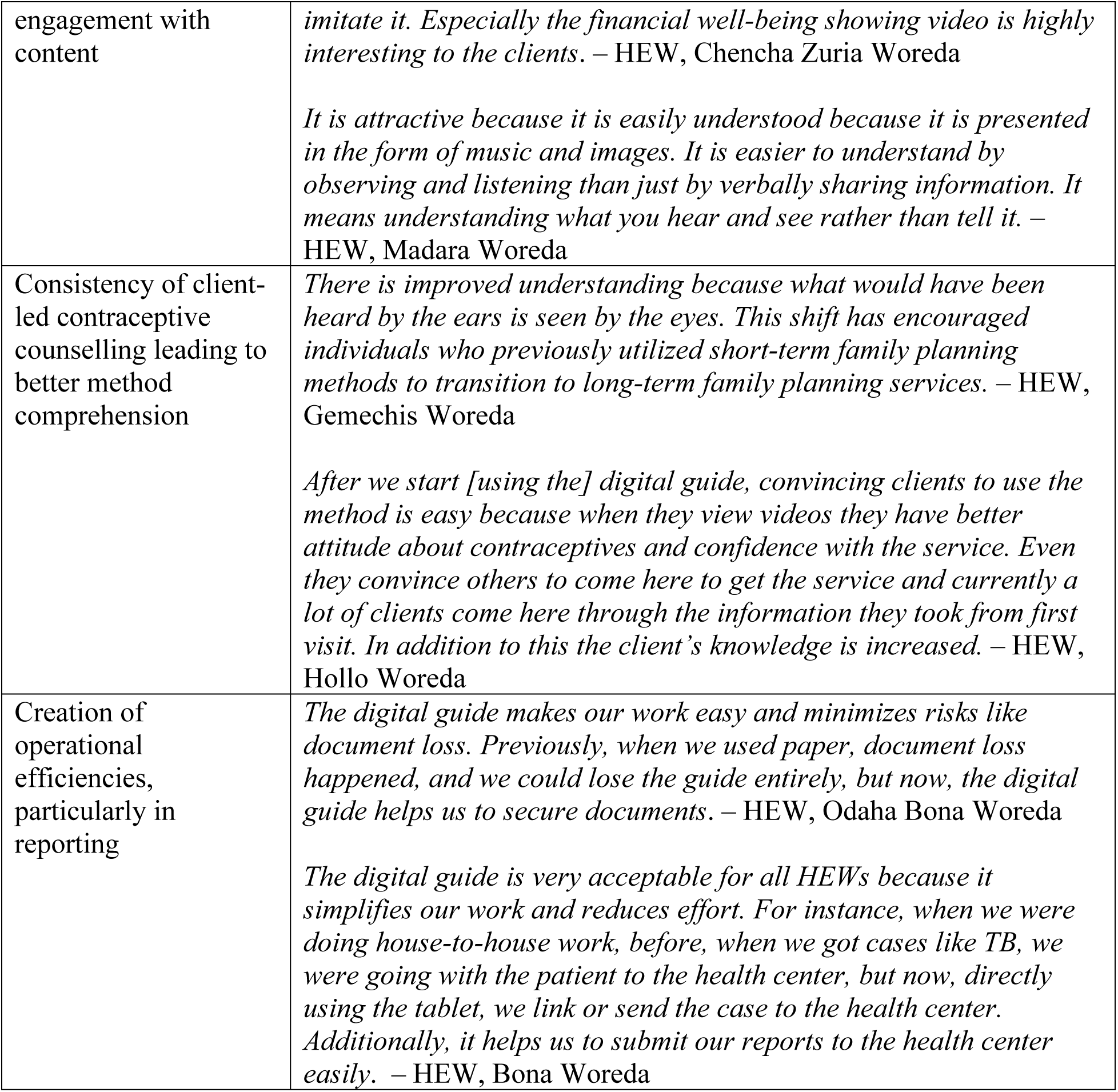
Advantages of using the Smart Start digital tool based on KIIs with HEWs.

HEWs commonly cited certain barriers to effective use of the digital counselling tool. Device-related limitations were most common, including glitches in the video playback, limited internet connectivity, and lack of power to the device. These limitations led HEWs in some instances to use the two versions of the tool concurrently. Use of the tool was also heavily influenced by HEWs’ own levels of digital literacy. Some HEWs were so unfamiliar with digital technology that they found using the digital guide to be insurmountable. Many HEWs said that one-off training wasn’t sufficient to enable successful use of the digital tool and requested further on-the-job training.

> *“During tablet use, issues such as system busyness and network problems hindering data synchronization occur. Additionally, the tablet may not charge properly, and occasionally, opening the tablet presents challenges.”* – HEW, Bona Woreda
>
> *“For me, paper-based guide is better than the digital guide. Because formerly I didn’t use an android phone in my life and when I start to use the digital guide, using the tablet is very difficult for me. Even when I was a student I used paper and I didn’t use a computer. The only time I used a computer was at the time of COC prior to using a digital guide.”* – HEW, Chencha Zuria Woreda

### Site-level productivity

Analysis of Health Management Information System (HMIS) data showed similar site-level productivity between paper-based and digital sites prior to the introduction of the digital tool, at around 26-27 contraceptive client visits per month among AGYW aged 15-24 (Figure 3 and Table 6). Contraceptive visits per site were relatively stable across both groups over the period prior to the introduction of the digital tool. During the introduction of the digital tool, productivity in both digital and paper-based sites decreased. In digital sites, this decrease was likely due to provider orientation to the new approach and training. In paper-based sites, the re-direction of technical assistance to digital sites may have resulted in reduced attention to implementation, alongside other programmatic changes occurring at the same time.

**Figure 3:**
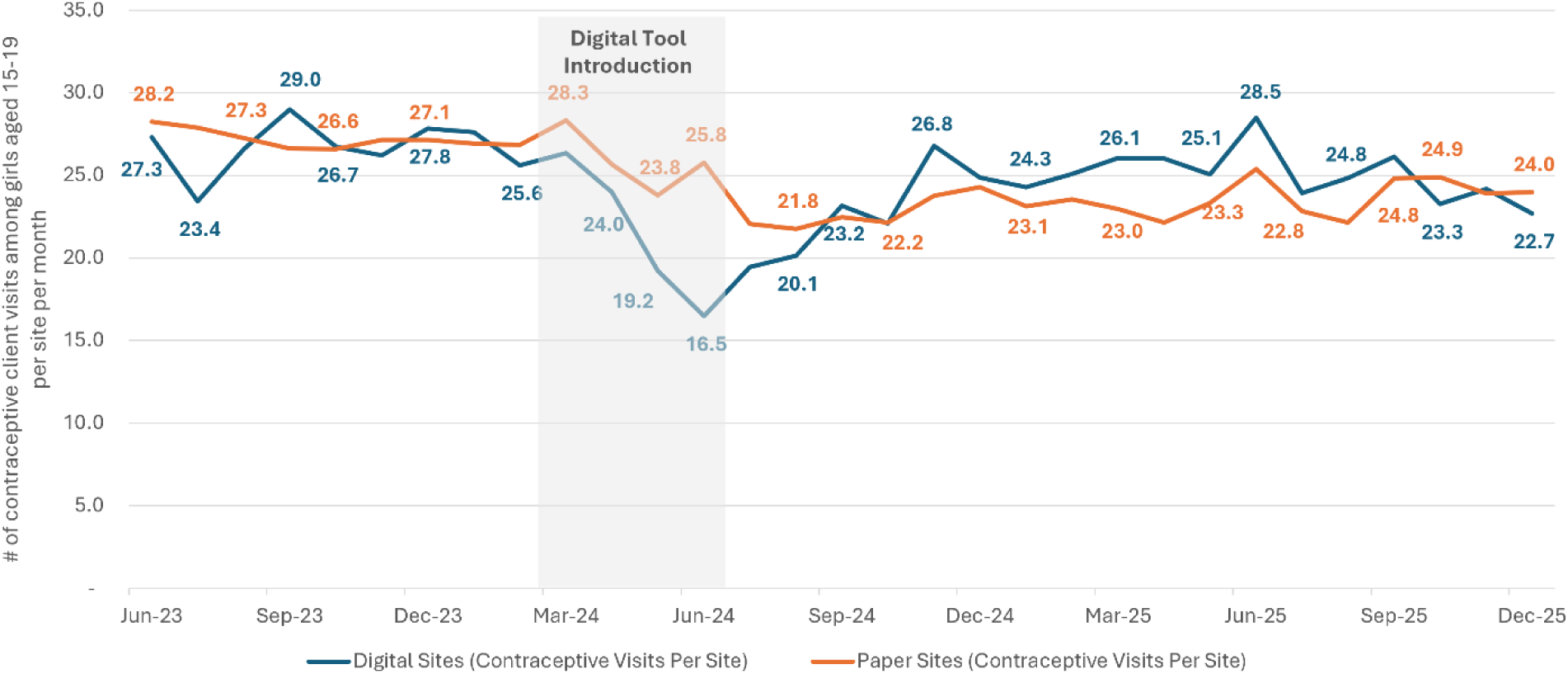
Contraceptive visits per site per month comparison between sites using digital and paper-based Smart Start counseling tool.

**Table 6:** Average contraceptive client visits per month per site comparison between sites using digital and paper-based Smart Start counseling tool.

|  | Before | Before | Before | During | After | After | After | After | After | After |
| --- | --- | --- | --- | --- | --- | --- | --- | --- | --- | --- |
|  | Q3 2023 | Q4 2023 | Q1 2024 | Q2 2024 | Q3 2024 | Q4 2024 | Q1 2025 | Q2 2025 | Q3 2025 | Q4 2025 |
| Digital Sites | 26.3 | 26.9 | 26.5 | 19.9 | 20.9 | 24.6 | 25.1 | 26.5 | 25.0 | 23.4 |
| Paper Sites | 27.3 | 27.0 | 27.4 | 25.1 | 22.1 | 23.4 | 23.2 | 23.6 | 23.3 | 24.3 |

After the initial transition period, digital sites showed some recovery in productivity, while paper-based sites remain at a slightly lower level. Digital site productivity then declined in late 2025. Overall, the introduction of the digital tool did not lead to sustained increases in service volume over the initial implementation phase compared to pre-intervention levels.

### Cost-efficiency

Comparison of unit cost per Smart Start session delivered shows higher cost for the digital tool of $10.94 compared to $1.94 for the paper-based tool (Table 7). The highest portion of the unit cost per session for the digital tool comes from training costs (comprising $8.80 of the total unit cost). Costs for development / maintenance and tool implementation were also higher for the digital tool. Given that the unit cost per session for the paper-based tool represents an institutionalized model (with training fully integrated within government-led training processes), we also sought to model out the prospective cost per session for the digital tool should it be similarly institutionalized. In doing so, the projected cost per session for the digital tool is reduced to $3.65, acknowledging that other components of the cost may be further reduced in a fully institutionalized model.

**Table 7:** Unit cost per Smart Start session delivered comparison between sites using digital and paper-based Smart Start counseling tool.

| Cost Component | Paper Based | Digital – Pilot | Digital –Training Institutionalized |
| --- | --- | --- | --- |
| Development and Maintenance | \$0.29 | \$0.50 | \$0.50 |
| Tool Implementation | \$0.12 | \$0.86 | \$0.86 |
| Training | \$0.76 | \$8.80 | \$1.51 |
| Counselor and client time | \$0.78 | \$0.78 | \$0.78 |
| **Total** | **\$1.94** | **\$10.94** | **\$3.65** |

## DISCUSSION

This study provides important evidence on the comparative experiential and counselling outcomes of digital versus paper-based counselling tools designed to support sexual and reproductive health among married AGYW in Ethiopia. Although both modalities supported high contraceptive uptake, the digitized Smart Start tool demonstrated advantages in counselling quality, client engagement, and informed decision-making. At the same time, operational and contextual barriers limited sustained use of the digital tool, underscoring the importance of context-sensitive approaches to digital integration within health systems.

One of the clearest advantages of the digital tool was improved counselling quality. Higher MII Plus scores among clients receiving digital counselling likely reflect the tool’s structured and standardized format, which appeared to support providers in consistently delivering complete information. Qualitative findings reinforced this interpretation, with HEWs reporting that the platform strengthened client-centered counselling and ensured key messages were systematically covered. These findings suggest that digital decision-support tools may reduce variability in provider performance and improve adherence to counselling protocols at scale, consistent with broader digital health evidence.^10,11^ Because MII Plus components are associated with contraceptive continuation, improved counselling quality may contribute not only to uptake but also to sustained contraceptive use over time.^12^

Digital counselling was also associated with more positive client experiences across several dimensions of care quality. Multimedia and interactive features appeared to shift counselling from a provider-directed interaction toward a more participatory and client-centered experience, improving engagement, comprehension, and trust, particularly among low-literacy populations. These findings suggest that the benefits of digital counselling extend beyond immediate service outcomes to include important changes in how counselling is delivered and experienced.

From the provider perspective, the digital tool offered operational advantages, including simplified workflows, improved data management, and stronger integration across services. HEWs reported that the platform facilitated referrals, reporting, and continuity of care while making counselling easier to conduct. These findings position the digital tool not only as a counselling aid but also as a broader health system strengthening intervention that may improve standardization, data visibility, and service coordination.

Importantly, the findings suggest that digital interventions may first influence processes, provider behaviors, and client experiences before measurable changes in service delivery outcomes emerge. HEWs described shifts in how they approached counselling, engaged clients, and used information to guide service delivery. Similarly, improvements in client experience and informed choice may represent early indicators of transformation that precede observable changes in utilization or productivity. Contraceptive uptake is influenced by multiple external factors, including population saturation, structural health system constraints, and demand-side barriers beyond the point of care.^13^ Consequently, the absence of immediate productivity gains should not necessarily be interpreted as limited effectiveness.

The transition from paper-based to digital counselling also presented implementation challenges. Connectivity limitations, hardware issues, and differences in provider digital literacy occasionally disrupted service delivery and affected usability. These findings suggest that constraints to scaling digital counselling may lie less in the intervention itself than in the enabling environment, including infrastructure, device reliability, and provider readiness.

HMIS service delivery data showed a temporary decline in productivity following digital tool introduction, likely reflecting the learning curve associated with adopting new technology. Although productivity recovered over time, it did not consistently exceed pre-intervention levels during the study period. However, traditional productivity metrics may not fully capture the broader value of digital interventions, particularly during early implementation when gains are more likely to occur in counselling quality, provider practices, client engagement, and data use rather than immediate increases in service volume.

Together, these findings highlight the importance of adequate transition periods, ongoing training, supportive supervision, and technical support during digital implementation. They also underscore the need for evaluation frameworks that capture intermediate process and quality improvements alongside longer-term service outcomes. Overall, the findings suggest that digital counselling tools can strengthen the quality and experience of contraceptive counselling, though achieving measurable service delivery gains may depend on sustained implementation support and broader health system readiness.

The digital counselling tool was associated with higher implementation costs than the paper-based model, though these findings should be interpreted considering differing stages of program maturity. The digital platform was implemented during a pilot phase requiring substantial upfront investments in software development, training, technical support, and system adaptation, whereas the paper-based model was already institutionalized within routine health system structures. Thus, cost differences likely reflect both the early-stage nature of digital implementation and the fixed investments required to establish digital systems. As digital tools become integrated into routine service delivery and implementation reaches scale, unit costs may decline through efficiencies in training, supervision, reporting, and data management. These findings highlight the importance of interpreting digital health costs dynamically across the implementation lifecycle rather than through short-term pilot comparisons alone.

### Conclusion

In conclusion, the digitalized Smart Start counselling tool demonstrated important advantages over the paper-based approach in improving counselling quality, client engagement, and informed choice among married AGYW in Ethiopia. Although measurable gains in service productivity were not apparent during the study period, findings suggest that digital tools may first drive changes in provider practices, client experience, and service quality before broader service delivery effects emerge. Successful scale-up of digital counselling will therefore depend not only on the effectiveness of the tool itself, but also on sustained investments in provider capacity, digital infrastructure, technical support, and health system readiness.

## MATERIALS AND METHODS

### Study Design

The study employed a cross-sectional mixed methods design where data was collected from health facilities implementing the digital and paper-based counselling tools. The study involved concurrent quantitative and qualitative data collection. Quantitative data entailed data from CEIs with AGYW who had received services from the health facilities and routine aggregate statistics on AGYW client contraceptive visits from the Ethiopia HMIS. Qualitative data involved key informant interviews (KIIs) with HEWs and midwives. Cost comparison drew financial and programmatic inputs from PSI and government records.

### Study Settings

The study was conducted in Sidama, Amhara, Oromia and South Ethiopia regions where the Smart Start intervention is implemented. Data was drawn from nine woredas, constituting 275 health posts and 55 health facilities where the Smart Start counseling tool was being implemented. Health posts and health centers are the lowest level of health facilities in the Ethiopian health system. Within these nine woredas, CEIs were conducted in a purposive sample of 32 health posts and 11 health centers. The paper-based tool facility sample included 16 health posts and eight health centers and the digital tool facility sample included 16 health posts and three health centers. HMIS data was analyzed only from the 275 health posts included in the overall sample (as this is the primary service delivery unit for Smart Start counseling).

### Sample size estimation

The sample size for CEIs was estimated using the formula for two independent proportions using method information index plus (MII Plus) as the primary estimator. MII Plus was employed to demonstrate the difference in the comprehensiveness of contraceptive counselling between clients from sites where digital and paper-based tools were used. We hypothesized that facilities where digital tools were employed would yield a higher proportion of clients responding affirmatively to the four MII Plus questions, better client experiences of the provider-client interaction, and better counselling outcomes. The sample was estimated to detect a 10-percentage point difference in MII Plus in favor of the digital sites and based on an average of 77% from three previous rounds of CEIs conducted for the Smart Start intervention where paper-based tools were used. A sample involving more than 300 clients was deemed adequate to yield 80% power to detect the 10% difference of MII Plus between the two groups. For the KIIs, the minimum number needed to attain code and thematic saturation was guided by the recommendations by Hennink et al (2017)^14^ who suggest between 16-24 interviews. For this study, 18 KIIs were conducted, which is within the recommended range. The facility sample for HMIS was purposely selected to match the facilities where clients were sampled for the CEIs. Costing data was analyzed at an aggregate level across all facilities implementing the paper-based and digital tools respectively.

### Participant and data collection procedures

AGYW clients who participated in CEIs were recruited using a systematic sampling approach based on daily attendance visits at the health facilities. Clients were eligible for participation if they were 15-24 years, were ever married, and had received contraceptive counselling and services from the selected health facilities on the duration of the survey. Trained female enumerators waited outside service delivery points where SRH services were provided, approached and screened women for eligibility using a recruitment script. Enumerators performed consenting procedures and conducted interviews in private spaces within the health facility. A quota of 10 clients was allocated per facility, and a skipping pattern was adopted for each facility based on previous caseloads for AGYW clients. Data was collected using an interviewer-administered survey tool customized on Survey CTO using the android application. Interviews took between 20-30 minutes and were conducted in English, Amharic, Afaan Oromo or Sidamo. Data was collected concurrently for the digital and paper-based group. Health workers who participated in KIIs were selected using purposive sampling ensuring representation from different woredas across the four regions. KIIs were conducted only with providers from sites utilizing the digital counselling tool (though prior to the digital tool introduction these providers had been using the paper-based tool). Trained qualitative researchers approached selected health workers, confirmed eligibility, and conducted consenting procedures prior to performing the interview within the health facility. Providers were eligible if they had been working at the facility for the six months prior to the study, were trained on and had been using the digital tools. KIIs took between 40 and 60 minutes, and all sessions were audio recorded. Monthly data on the aggregate numbers of clients 15-24 years who received contraceptive services from the period June 2023 to December 2025, as generated from service registers, was drawn from HMIS using the District Health Information Software 2 (DHIS2) platform into an Excel database. This period represented one year prior and 19 months after the introduction of the digital tool. We created an Excel-based costing tool to input data to generate a cost comparison between the digital and paper-based tools. Data for this costing component were generated from a variety of sources including PSI financial records, site-level government cost surveys, and program data.

### Measures

The CEI survey tool contained questions to capture participant social and demographic characteristics, MII Plus, clients’ perceptions of the counselling tool and experiences with the provider-client interaction and services. Socio-demographic questions captured location (urban vs rural), age, schooling status, highest educational level, religion, parity and exposure to Smart Start. MII Plus was measured using the standard four questions with ‘yes’ or ‘no’ response options.^15^ Clients’ perceptions of the counselling tools used were measured with a Likert scale with statements eliciting three response options: ‘agree’, ‘indifferent’ and ‘disagree’. The clients’ experiences of the provider-client interaction during the counselling session were measured using a scale involving statements with participants expected to respond with five response options: ‘strongly agree’, ‘agree’, neutral’, ‘disagree’ or ‘strongly disagree’. KIIs were moderated using facilitation guides with questions to assess providers’ experiences with using the digitalized tool, perception of the support provided during implementation, challenges encountered and recommendations. HMIS data captured aggregate monthly contraceptive client visits (which include a combination of new and repeat clients) by facility. Costing measures were generated using financial inputs related to tool development and maintenance, training development and delivery, printing and distribution (paper-based tool only), tablet purchase and running costs (digital tool only), and health workforce associated costs (for example HEW annual salaries). These were combined with programmatic inputs including assumptions on frequency of updates for tools, training, and equipment replacement; the proportion of time for equipment use on Smart Start counseling (digital tool only); and health worker and client interaction metrics (number of health workers implementing the tool, average time with a client, average number of clients seen per year per counselor, and others). The final model output represented a cost per Smart Start counseling session delivered.

### Data analysis

For CEIs, descriptive analysis was employed to generate summaries of the socio-demographic, MII Plus, perceptions of the tools and client experience questions. MII Plus was measured as a composite binary variable computed using the approach by Jain et al where clients who responded with a ‘yes’ to all four questions were coded as 1. Clients responding with ‘no’ to any of the four questions were coded as 2.^15^ Clients’ responses on the perception of the tools used were reduced to a binary variable by collapsing the ‘indifferent’ and ‘disagree’ responses as a ‘no’ and retaining the ‘agree’ response as ‘yes’. Similarly, the five response options on provider-client interactions were reduced so that ‘strongly agree’ and ‘agree’ were coded as ‘agree’ and the other response options coded ‘not agree’. Tests of equality (chi square test or Fischer exact test) were used to determine whether the differences between the digital and paper-based groups were significant. Audio files from the KIIs were transcribed verbatim, proofread to check and correct any errors and organized in Dedoose. Thematic analysis was conducted using both inductive and deductive approaches employing a codebook developed following the structure of the core and probing questions in the facilitation guides. Analysis of monthly aggregate service data from HMIS entailed descriptive and trend analysis. In Excel, facility-level monthly client visit numbers were aggregated within two groups representing paper-based and digital tool implementation. For each group respectively, monthly aggregate client visit numbers for the entire group were then divided by the total number of health posts with that respective implementation group (183 for the paper-based group and 92 for the digital group). The resulting monthly contraceptive client visits per site per group were visualized in tabular and graphical format. The costing tool assessed costs for each version of the tool (digital and paper-based) according to categories related to a) tool development and maintenance, b) tool implementation costs, c) training costs, and d) counselor and client time costs. For categories a through c, we aggregated all costs within the category and then divided by total number of HEWs utilizing the tool to come up with a cost per counselor per year and then further divided by the number of clients seen by each HEW per year to come to a cost per session. For category d, these costs were aggregated and only divided by number of clients seen by HEW per year. The cost per session for all categories were then combined to arrive at a total cost per Smart Start session delivered for each tool. As the model was particularly sensitive to scale (the number of HEWs implementing each tool respectively), we maintained this number as static between groups to promote appropriate comparison even though the paper-based tool is being implemented currently at a much larger scale.

### Ethics Statement

Ethics approvals were obtained for the CEIs and qualitative data collection from the Ethiopian Midwives Association Institutional Review Board (approval #EMwA-IRB-SOP/007/10-24) and the Population Services International Research Ethics Board (approval #1778/2023). Written informed consent was granted by all CEI and qualitative participants prior to their participation. As all CEI participants were married, they were considered emancipated minors and able to provide informed consent directly for participation. A waiver of parental consent for adolescent girls aged 15-17 years was granted by the approving institutions.

### Study Limitations

This study contributes to the growing evidence base about use of digital tools to support providers during service provision by improving the provider-client decision making process. The study employed a mixed-methods approach triangulating between different data sources, but not without limitations. First, the cross-sectional design used does not support drawing conclusions about whether the differences observed were specifically attributable to the digital tools or due to unstudied background characteristics between the clients, providers and health facilities. Second, the study relied on self-reports from clients and providers which are susceptible to social desirability biases. Third, data was collected within the context of an existing program using a novel counselling approach that has proven effective at increasing contraceptive use indicators and outcomes. The comparisons were made between digital and paper-versions of the Smart Start tool instead of comparison with standard counselling employed in the Ethiopia health system. The similarities between these tools may have contributed to the failure to observe differences as both tools yielded comparable client experiences and perceptions across multiple areas.

## DATA AVAILABILITY

Quantitative datasets used and analyzed for this study (including CEI, HMIS, and costing summaries) are publicly available via figshare (DOI: 10.6084/m9.figshare.32253591; 10.6084/m9.figshare.32253621; 10.6084/m9.figshare.32253630). Qualitative data from KIIs is not shared publicly given these data cannot be completely de-identified.

## ACKNOWLEDGMENTS

The authors wish to acknowledge support from the team supporting implementation of this project by Population Services International Ethiopia as well as the Ethiopian Federal Ministry of Health and Regional and Zonal Health Bureaus who supported conceptualization and implementation of this work.

